# Machine Learning-Based Mortality Prediction in Critically Ill Patients with Hypertension: Comparative Analysis, Fairness, and Interpretability

**DOI:** 10.1101/2025.04.05.25325307

**Authors:** Shenghan Zhang, Sirui Ding, Zidu Xu, Jiancheng Ye

## Abstract

**Background:** Hypertension is a leading global health concern, significantly contributing to cardiovascular, cerebrovascular, and renal diseases. In critically ill patients, hypertension poses increased risks of complications and mortality. Early and accurate mortality prediction in this population is essential for timely intervention and improved outcomes. Machine learning (ML) and deep learning (DL) approaches offer promising solutions by leveraging high-dimensional electronic health record (EHR) data.

**Objective:** To develop and evaluate ML and DL models for predicting in-hospital mortality in hypertensive patients using the MIMIC-IV critical care dataset, and to assess the fairness and interpretability of the models.

**Methods:** We developed four ML models—gradient boosting machine (GBM), logistic regression, support vector machine (SVM), and random forest—and two DL models— multilayer perceptron (MLP) and long short-term memory (LSTM). A comprehensive set of features, including demographics, lab values, vital signs, comorbidities, and ICU-specific variables, were extracted or engineered. Models were trained using 5-fold cross-validation and evaluated on a separate test set. Feature importance was analyzed using SHapley Additive exPlanations (SHAP) values, and fairness was assessed using demographic parity difference (DPD) and equalized odds difference (EOD), with and without the application of debiasing techniques.

**Results:** The GBM model outperformed all other models, with an AUC-ROC score of 96.3%, accuracy of 89.4%, sensitivity of 87.8%, specificity of 90.7%, and F1 score of 89.2%. Key features contributing to mortality prediction included Glasgow Coma Scale (GCS) scores, Braden Scale scores, blood urea nitrogen, age, red cell distribution width (RDW), bicarbonate, and lactate levels. Fairness analysis revealed that models trained on the top 30 most important features demonstrated lower DPD and EOD, suggesting reduced bias. Debiasing methods improved fairness in models trained with all features but had limited effects on models using the top 30 features.

**Conclusions:** ML models show strong potential for mortality prediction in critically ill hypertensive patients. Feature selection not only enhances interpretability and reduces computational complexity but may also contribute to improved model fairness. These findings support the integration of interpretable and equitable AI tools in critical care settings to assist with clinical decision-making.

## INTRODUCTION

Hypertension, commonly known as high blood pressure, is one of the most prevalent chronic conditions globally and a leading contributor to morbidity and mortality. According to the World Health Organization, approximately 1.13 billion people worldwide suffer from hypertension, with fewer than 20% achieving adequate blood pressure control.[1] The condition significantly increases the risk of cardiovascular disease, stroke, and chronic kidney disease, often resulting in poor health outcomes and elevated healthcare costs.[2] Despite advances in pharmacological treatments and lifestyle interventions,[3, 4] hypertension remains a major public health challenge and a key contributor to premature mortality.[5]

In critically ill patients, the presence of hypertension further complicates clinical trajectories, especially in intensive care unit (ICU) settings where rapid deterioration can occur.[6] Early identification of high-risk individuals among hypertensive patients admitted to the ICU is essential for timely and targeted interventions that could reduce mortality rates and optimize resource allocation.[7] However, traditional mortality prediction models, such as risk scoring systems, often rely on a narrow set of pre-selected variables and may not adequately capture the complex, nonlinear relationships and dynamic physiological changes present in critically ill populations. Recent advances in machine learning (ML) and deep learning (DL) have opened new avenues for clinical risk prediction by enabling the analysis of large-scale electronic health record (EHR) data with minimal prior assumptions.[8] These models are capable of learning complex patterns from high-dimensional data and can incorporate a wide range of clinical variables—including demographic data, comorbidities, laboratory test results, vital signs, and treatment information—into predictive frameworks.[9] Among the rich sources of EHR data available for research, the Medical Information Mart for Intensive Care IV (MIMIC-IV) database presents a comprehensive, publicly available resource containing detailed longitudinal data on ICU patients.

In this study, we utilize the MIMIC-IV dataset to develop and evaluate a suite of ML and DL models for predicting in-hospital mortality among patients diagnosed with hypertension. We explore six modeling approaches, including four machine learning algorithms—gradient boosting machine (GBM), logistic regression, support vector machine (SVM), and random forest—and two deep learning architectures—a multilayer perceptron (MLP) and long short-term memory (LSTM) network. We incorporate a broad set of features and introduce new variables related to ICU complications and patient conditions to enhance predictive performance.

Our work has three objectives: (1) to assess the comparative performance of these models in predicting mortality in critically ill patients with hypertension, (2) to identify key clinical features associated with increased mortality risk using explainable techniques such as SHAP (SHapley Additive exPlanations), and (3) to examine the fairness of model predictions across demographic groups while testing debiasing strategies. By integrating prediction accuracy, interpretability, and fairness evaluation, this study also aims to contribute to the growing body of evidence supporting the use of AI-driven models in clinical decision-making. Ultimately, our goal is to lay the groundwork for developing scalable, equitable, and actionable tools to assist clinicians in managing hypertensive patients in critical care settings.

## METHODS

### Dataset and Study Population

This study leverages the Medical Information Mart for Intensive Care IV (MIMIC-IV) dataset, which comprises detailed clinical data from patients admitted to critical care units at a large tertiary care hospital.[10] The dataset includes demographics, vital signs, laboratory results, medications, interventions, and outcomes for over 40,000 patients. MIMIC-IV is structured into modules corresponding to various hospital departments, allowing access to patient data at both hospital and ICU levels. Using unique patient identifiers, we integrated information across these modules to construct a comprehensive cohort. For this study, we focused on predicting mortality among patients diagnosed with hypertension. **Figure 1** illustrates the overall study pipeline. We identified all adult patients (aged 18 and older) with hypertension using ICD-9 and ICD-10 diagnosis codes. Mortality outcomes were determined based on hospital-level data. Patients with incomplete records or missing outcome data were excluded from the analysis.

**Figure 1.**
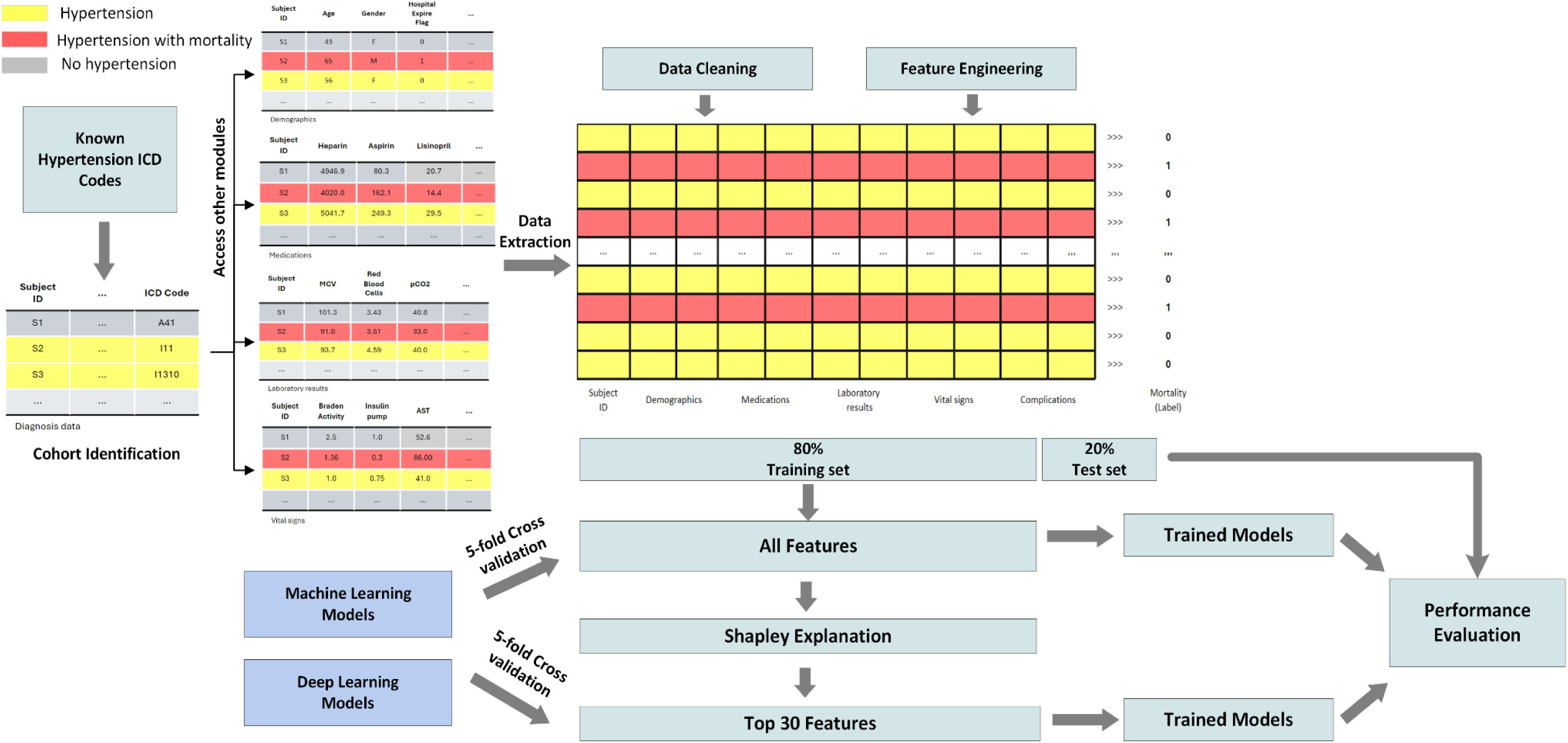
Overview of the hypertension mortality prediction pipeline.

### Data Preprocessing Pipeline

Data preprocessing was essential to ensure the quality and robustness of our machine learning models.[11] The pipeline included four major steps: (1) data extraction, (2) data cleaning, (3) feature engineering, and (4) feature selection. Given the breadth of conditions represented in MIMIC-IV, we first isolated patients with hypertension using relevant ICD-10 codes. ICD-9 codes were mapped to ICD-10 using the General Equivalence Mapping (GEM) tables provided by the Centers for Medicare & Medicaid Services (CMS), ensuring a unified coding system. Mortality status was derived from the hospital_expire_flag variable. Each patient was identified using a unique subject_id to extract information across modules. The features collected included (1) demographics (age, sex, self-reported race), (2) laboratory test results, (3) medication history, (4) vital signs, and (5) clinical complications.

To address data quality, we eliminated duplicate records by retaining the most recent entry per patient. Features with more than 80% missing data were discarded, and remaining missing values were imputed using the median. Patients with ambiguous demographic information (e.g., “unknown” or “other” for race) were excluded to minimize potential biases. Feature engineering involved creating new variables based on hospital and ICU-level data. We calculated the length of hospital stay (in days), and binary indicators were created for ICU admission and clinical complications (based on ICD-10 codes). Continuous variables were normalized to ensure comparability. Race was encoded using binary indicators for each category.

### Feature Selection and Model Explainability

Feature selection combined domain expertise with automated methods, including recursive feature elimination and feature importance rankings derived from preliminary models. Key predictors of mortality included age, sex, vital signs (e.g., systolic/diastolic blood pressure, heart rate), laboratory values (e.g., creatinine, potassium), and treatment indicators (e.g., antihypertensive use). To enhance interpretability, we used SHapley Additive exPlanations (SHAP, version 0.47.1) to compute feature importance for both machine learning and deep learning models.[12] SHAP values quantify the marginal contribution of each feature to a prediction, averaged across all instances. Features were ranked by their average SHAP value, and the top 30 were retained for model development.

### Model Development

To comprehensively evaluate model performance, we developed and trained four machine learning (ML) models and two deep learning (DL) models on our hypertension patient cohorts. The ML models included logistic regression, random forest, support vector machines (SVM), and gradient boosting machines (GBM), selected for their ability to handle the complexity of healthcare data and their varying degrees of interpretability.[13] Logistic Regression is a linear model widely used in medical research due to its simplicity and high interpretability, making it a common baseline for binary classification tasks. Random Forest is an ensemble learning method that aggregates multiple decision trees to model non-linear relationships and interactions between features, improving prediction robustness. SVM classify outcomes by identifying the optimal hyperplane that separates data points in a high-dimensional space. SVMs are particularly effective when the relationship between predictors and outcomes is non-linear. GBM are another ensemble method that iteratively combines weak learners, often decision trees, to minimize a loss function using gradient descent. GBMs are known for strong predictive performance and flexibility. These four ML models have been widely applied in the literature for mortality prediction using EHR data and are well-regarded for balancing accuracy and interpretability.[14]

Additionally, we implemented two deep learning models—Multi-Layer Perceptrons (MLP) and Long Short-Term Memory (LSTM) networks—both commonly applied in clinical outcome prediction tasks.[15] MLP are feedforward neural networks comprising multiple layers of interconnected neurons. We implemented a standard three-layer MLP, with the input layer matching the number of patient features, a hidden layer containing 128 neurons, and an output layer with two neurons and softmax activation. LSTM networks is a specialized type of recurrent neural network, are designed to capture temporal dependencies in sequential data. This makes them well-suited for modeling longitudinal patient data. We developed an LSTM model with an input dimension based on patient features and a hidden state size of 128, followed by a softmax-activated output layer with two neurons.

All models were developed using Python 3.8.19. ML models (logistic regression, random forest, SVM) were implemented using the scikit-learn library (v1.3.0), and GBM was implemented using the XGBoost library (v2.0.3). DL models were developed using PyTorch (v2.3.0), trained for 500 epochs using the Adam optimizer (learning rate = 1e-3, batch size = 1024).

### Model Fairness and Bias Mitigation

Fairness is a critical consideration when developing machine learning models for clinical use. Ensuring fairness entails minimizing algorithmic bias across demographic groups, such as sex or race.[16] In this study, we focused on identifying and mitigating biases across sex subgroups. Biases can generally be classified into two categories: data-level bias and model-level bias. To address both, we implemented three types of debiasing strategies described in recent literature[17]: 1) Correlation Removal, a pre-processing technique, eliminates features that are highly correlated with sensitive demographic variables to reduce data-level bias; 2) Reduction, an in-processing method, introduces a fairness-related penalty term in the model’s loss function during training to discourage biased predictions; 3) Threshold Optimization, a post-processing method, adjusts the model’s decision threshold to improve fairness metrics without retraining.

We used the Fairlearn library (v0.10.0) to implement the reduction and threshold optimization methods for ML models. Due to Fairlearn’s limited support for DL models, we manually implemented equivalent debiasing strategies, including a custom reduction term and correlation-based feature filtering.

### Model Evaluation and Statistical Analysis

The dataset was randomly partitioned into a training set (80%) and a test set (20%). We applied 5-fold cross-validation on the training set to tune hyperparameters and prevent overfitting. Final evaluations were conducted on the held-out test set to assess generalizability. To evaluate the utility of our feature selection strategy, models were trained using both the original extracted features and a reduced feature set filtered by SHAP values. Performance comparisons on the test set enabled assessment of the impact of feature selection on prediction accuracy. Model performance was assessed using standard metrics for clinical prediction tasks, including accuracy, sensitivity (recall), specificity (precision), F1 score, area under the receiver operating characteristic curve (AUC-ROC), and area under the precision-recall curve (AUPRC). Accuracy and AUC-ROC reflect overall model discriminative ability, while sensitivity, specificity, and F1 score provide insight into performance among individuals with mortality.

To compare patient characteristics between those with and without mortality, Welch’s t-tests were used for continuous variables and Chi-square tests for categorical variables. A p-value < 0.05 was considered statistically significant. For performance metrics, we computed 95% confidence intervals (CI) using the following approaches: 1) Normal approximation for cross-validation results; 2) Wald intervals for accuracy, sensitivity, and specificity on the test set; 3) Bootstrapping (1,000 iterations) for complex metrics, such as AUROC and F1 score. Model fairness was evaluated using two metrics: 1) Demographic Parity Difference (DPD): the difference in positive prediction rates between demographic groups; and 2) Equalized Odds Difference (EOD): the difference in true positive and false positive rates across groups. Bootstrapping (1,000 iterations) was used to calculate 95% CIs for both fairness metrics.

### Study Approval

We solely used publicly available MIMIC-IV data for this study.

## RESULTS

### Patient characteristics

**Table 1** demonstrates the characteristics of the patient cohort, including distributions for age, length of hospital stay, sex, race, and clinical complications. After applying the inclusion and exclusion criteria, we identified 88,084 patients diagnosed with hypertension. Among them, 3.9% (3,442 patients) died during their hospital stay. The cohort was composed of 48.6% females and 51.4% males, with a mean age of 48.9 years (standard deviation [SD] = 14.8 years). The average age of patients in the mortality group was significantly higher at 56.4 years (SD = 12.8), compared to 48.6 years (SD = 14.8) in the non-mortality group, suggesting that older patients face a greater risk of in-hospital mortality. This age difference was also reflected in the median values: 59 years (IQR: 48–67) in the mortality group versus 49 years (IQR: 39–60) in the non-mortality group. There was a substantially higher ICU stay rate among patients who died (87.0%) compared to those who survived (37.7%).

**Table 1.** Characteristics of the patients.

| Characteristics | Total (n=88084) | Mortality (n=3442) | Non-mortality (n=84642) | P-value |
| --- | --- | --- | --- | --- |
| **Sex** |  |  |  | 0.451 |
| Male | 45241 (51.4%) | 1790 (52.0%) | 43451 (51.3%) |  |
| Female | 42843 (48.6%) | 1652 (48.0%) | 41191 (48.7%) |  |
| **Age** |  |  |  |  |
| Mean (SD) | 48.93 (14.79) | 56.38 (12.78) | 48.63 (14.79) | <.001 |
| **Length of stay, days** |  |  |  |  |
| Median (IQR) | 6.9 (2.9-16.2) | 7.49 (2.7-16.6) | 6.9 (2.9-16.2) | <.001 |
| **Race** |  |  |  | <0.01 |
| American Indian | 175 (0.2%) | 2 (0.1%) | 173 (0.2%) |  |
| Asian | 2696 (3.1%) | 112 (3.3%) | 2584 (3.1%) |  |
| Black | 12299 (14.0%) | 257 (7.5%) | 12042 (14.2%) |  |
| Hispanic | 3831 (4.3%) | 61 (1.8%) | 3770 (4.5%) |  |
| White | 60425 (68.6%) | 2129 (61.9%) | 58296 (68.9%) |  |
| Others | 8658 (9.8%) | 881 (25.6%) | 7777 (9.2%) |  |
| **ICU stay** |  |  |  | <.001 |
| Yes | 34868 (39.6%) | 2993 (87.0%) | 31875 (37.7%) |  |
| No | 53216 (60.4%) | 449 (13.0%) | 52767 (62.3%) |  |
| **Complication** |  |  |  |  |
| Diabetes Mellitus | 51690 (58.7%) | 1585 (46.0%) | 50105 (59.2%) | 0.194 |
| Heart Failure | 43593 (49.5%) | 2193 (63.7%) | 41400 (48.9%) | <.001 |
| Arrhythmias | 8278 (9.4%) | 421 (12.2%) | 7857 (9.3%) | <.001 |
| Myocardial Infarction | 7591 (8.6%) | 526 (15.3%) | 7065 (8.3%) | <.001 |
| Chronic Obstructive Pulmonary Disease | 14424 (16.4%) | 788 (22.9%) | 13636 (16.1%) | <.001 |
| Asthma | 9889 (11.2%) | 177 (5.1%) | 9712 (11.5%) | <.001 |
| Pneumonia | 13085 (14.9%) | 970 (28.2%) | 12115 (14.3%) | <.001 |
| Stroke | 3397 (3.9%) | 278 (8.1%) | 3119 (3.7%) | <.001 |
| Dementia | 8427 (9.6%) | 379 (11.0%) | 8048 (9.5%) | <.001 |

### Model Performance Using All Extracted Features

We trained several models on the hypertension cohort and evaluated their performance using 5-fold cross-validation and a separate held-out test set. Performance was assessed using five metrics: Accuracy, Area Under the Receiver Operating Characteristic Curve (AUROC), Sensitivity (Recall), Specificity (Precision), and F1 Score. For each metric, we report the mean and the corresponding 95% confidence interval (CI), reflecting the variability across data partitions. Full cross-validation results are available in **Supplementary Table 1**. Overall, deep learning models outperformed traditional machine learning approaches, indicating their strength in capturing complex patterns. The Long Short-Term Memory (LSTM) network achieved the highest performance across all metrics, with accuracy of 93.9%, sensitivity of 94.3%, specificity of 93.5%, and an F1 score of 93.9%. Among machine learning models, logistic regression (LR) performed the worst, while gradient boosting machines (GBM) showed the best performance, particularly in accuracy, AUROC, and specificity.

To evaluate generalizability, we also assessed performance on the test set (**Table 2**). Notably, both MLP and LSTM models exhibited performance declines—3.2% and 5.5% decreases in accuracy, respectively—across all metrics, suggesting potential overfitting. In contrast, the machine learning models (LR, RF, SVM, and GBM) maintained stable performance, underscoring better generalizability and lower risk of overfitting. **Figure 2** displays the AUC-ROC and Precision-Recall (PR) curves. The ROC curves illustrate the trade-off between true positive rate and false positive rate, with GBM achieving the highest AUC (0.97), closely followed by LR, RF, and SVM (all with AUC = 0.96). The PR curves evaluate the balance between precision and recall, where GBM again outperformed other models (AUC = 0.97), followed by LR, RF, and SVM (0.96), LSTM (0.94), and MLP (0.93). These curves indicate that GBM consistently offers the best balance of sensitivity and precision for predicting mortality in hypertensive patients.

**Table 2.**
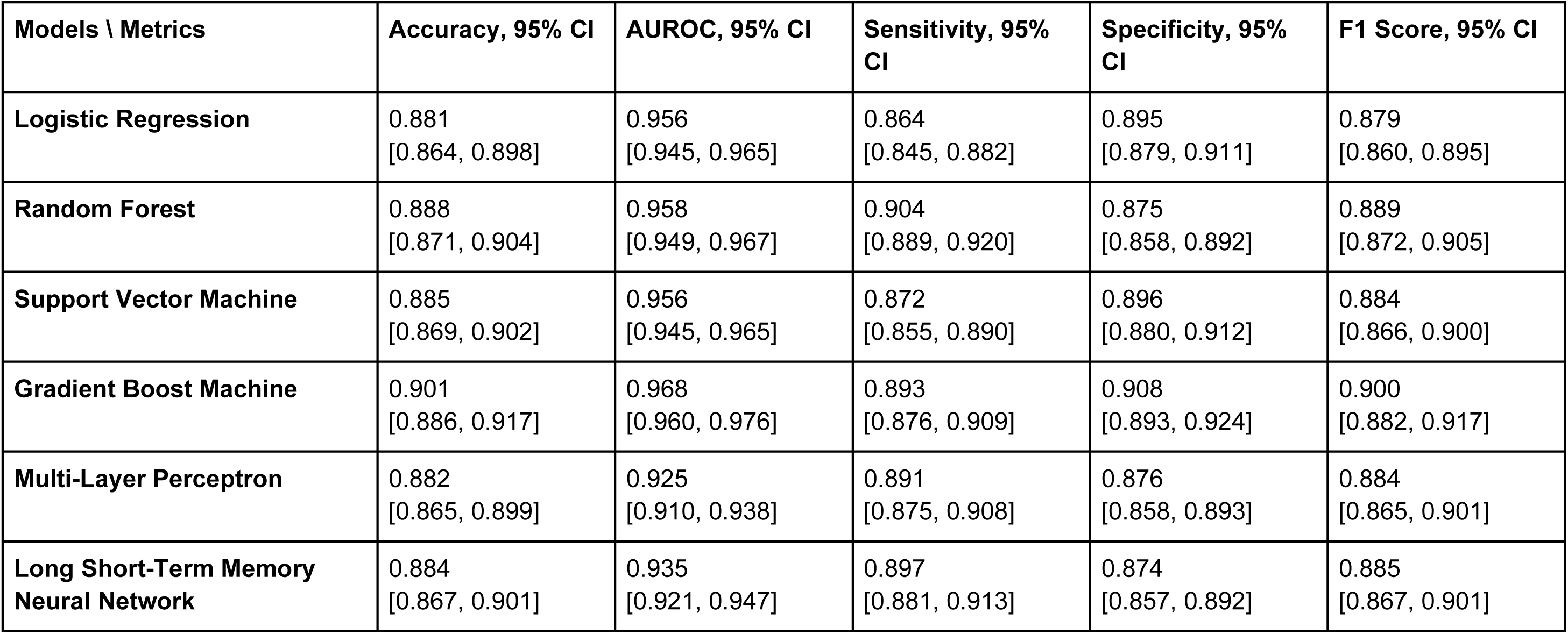
Model performance for hypertension mortality prediction on the test set using all features.

**Figure 2.**
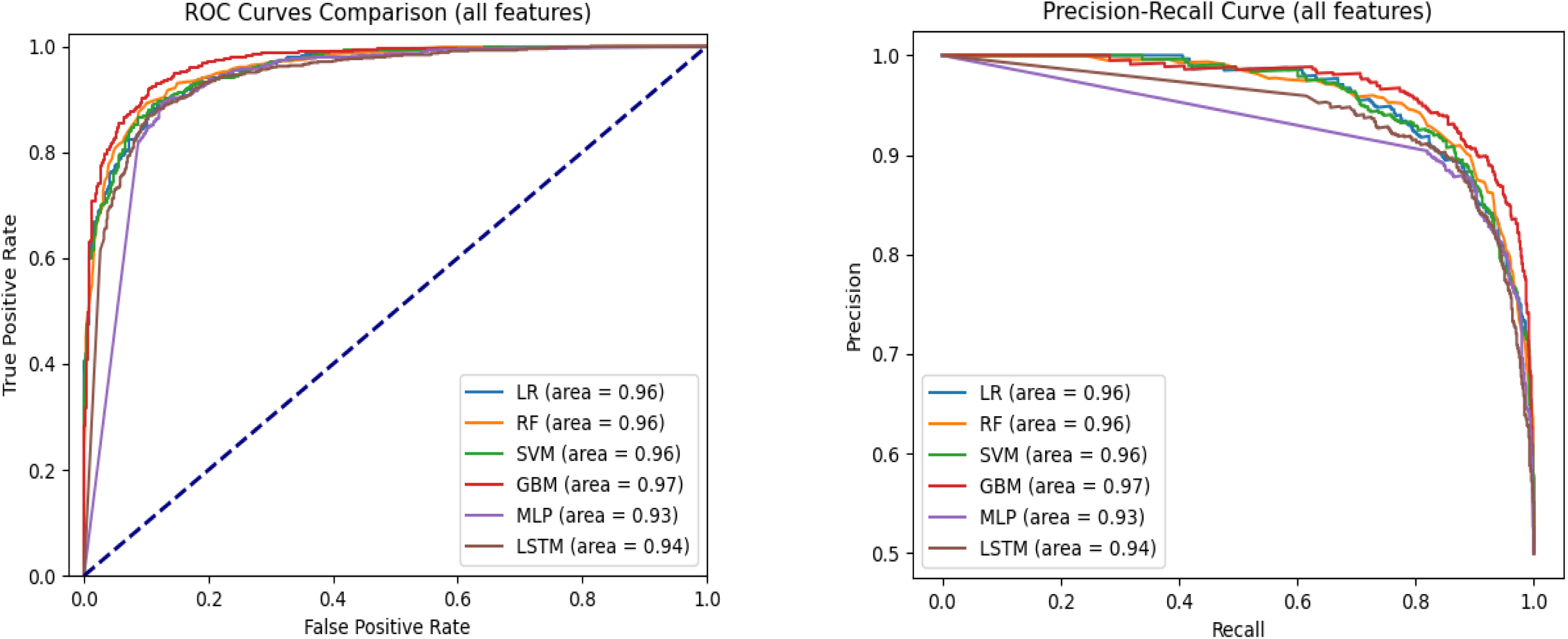
AUC-ROC and Precision-Recall curves comparing model performance for hypertension mortality prediction on the test set using all features.

### Feature Importance Analysis and Feature Selection

To address potential issues related to overfitting and model complexity, we conducted a feature importance analysis to guide feature selection. Using SHAP values, we quantified the importance of each feature across all models (LR, RF, SVM, GBM, MLP, and LSTM), based on the 5-fold cross-validation results. **Figure 3** presents the top 30 features ranked by average SHAP values. The most influential predictors were Glasgow Coma Scale (GCS) eye-opening response, emergency admission type, anchor age, Braden mobility score, and urea nitrogen level. These features consistently ranked highly across models and are likely key indicators of mortality risk in patients with hypertension. To reduce model complexity and mitigate overfitting, we limited the feature set to these top 30 most important variables for subsequent modeling.

**Figure 3.**
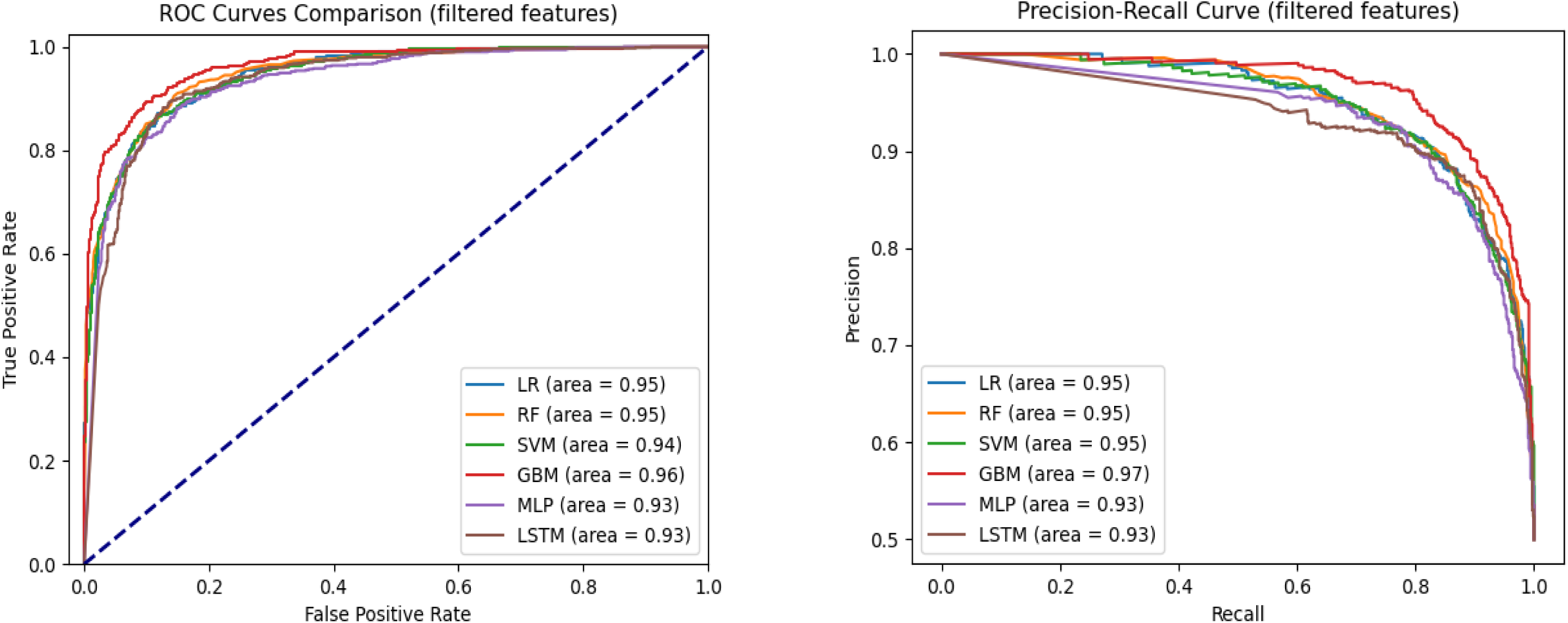
AUC-ROC and Precision-Recall curves comparing model performance for hypertension mortality prediction on the test set using top 30 features.

### Model Performance with Selected Important Features

After selecting the top 30 most important features, we retrained and fine-tuned all models using 5-fold cross-validation. These validated models were then evaluated on a held-out 20% test set to assess their generalizability to unseen data. Across models, performance differences between using all features versus the top 30 were minimal—typically under 0.2%—indicating stable model behavior with no signs of overfitting.

Model performance patterns observed with all features were largely preserved when using only the top 30 features during cross-validation. Among all models, the Gradient Boosting Machine (GBM) achieved the highest performance, with an AUROC of 96.0% (95% CI: 95.6%, 96.4%) and an F1 score of 89.1% (95% CI: 88.2%, 90.0%). Both the Multi-Layer Perceptron (MLP) and Long Short-Term Memory (LSTM) models outperformed most traditional machine learning models, except for GBM. The Logistic Regression (LR) model had the lowest performance across accuracy (86.1%), sensitivity (84.0%), and F1 score (85.8%). The MLP model reported the lowest AUROC (93.2%), while the Random Forest (RF) had the lowest specificity (86.9%). Full cross-validation metrics with 95% confidence intervals are provided in **Supplementary Table 2**.

### Test Set Evaluation

On the test set, the LR model achieved an accuracy of 86.9%, AUROC of 94.6%, sensitivity of 85.2%, specificity of 88.3%, and F1 score of 86.7%. The RF model slightly outperformed LR, with an accuracy of 87.4%, AUROC of 94.8%, sensitivity of 88.8%, specificity of 86.4%, and F1 score of 87.6%. The Support Vector Machine (SVM) also demonstrated strong performance: accuracy of 87.2%, AUROC of 94.5%, sensitivity of 85.3%, specificity of 88.7%, and F1 score of 87.0%. GBM again delivered the best performance across most metrics, achieving an accuracy of 89.4% and AUROC of 96.3%, with sensitivity and specificity of 87.8% and 90.7%, respectively, and an F1 score of 89.2%. In contrast, the deep learning models saw a slight decline in test performance. The MLP achieved 86.0% accuracy, 93.2% AUROC, sensitivity of 86.4%, specificity of 85.7%, and an F1 score of 86.0%. LSTM performed better than MLP, with 87.4% accuracy, 92.7% AUROC, sensitivity of 87.5%, specificity of 87.3%, and an F1 score of 87.4%. These results suggest that while deep learning models performed competitively in cross-validation, they did not surpass traditional machine learning models on the held-out test set. Test set results are summarized in **Table 3**.

**Table 3.**
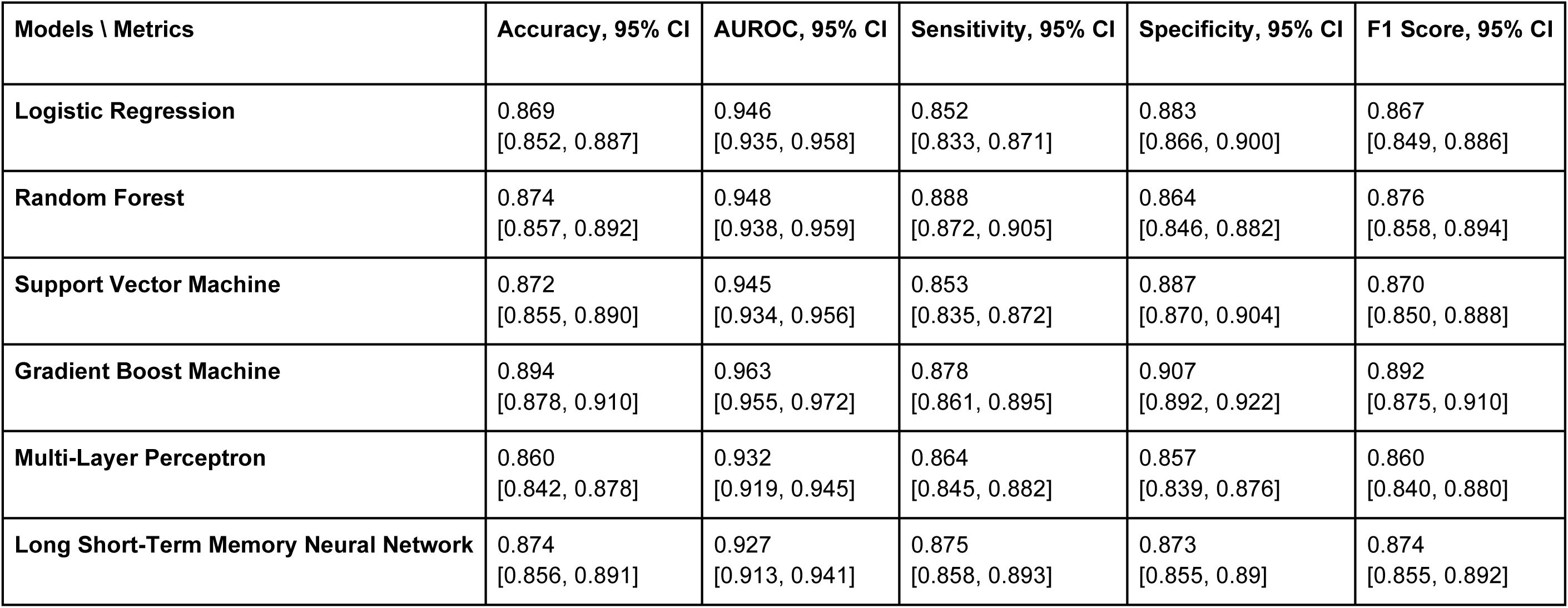
Summary of model performance for hypertension mortality prediction on the test set using the top 30 selected features.

| Models \ Metrics | Accuracy, 95% CI | AUROC, 95% CI | Sensitivity, 95% CI | Specificity, 95% CI | F1 Score, 95% CI |
| --- | --- | --- | --- | --- | --- |
| **Logistic Regression** | 0.869<br>[0.852, 0.887] | 0.946<br>[0.935, 0.958] | 0.852<br>[0.833, 0.871] | 0.883<br>[0.866, 0.900] | 0.867<br>[0.849, 0.886] |
| **Random Forest** | 0.874<br>[0.857, 0.892] | 0.948<br>[0.938, 0.959] | 0.888<br>[0.872, 0.905] | 0.864<br>[0.846, 0.882] | 0.876<br>[0.858, 0.894] |
| **Support Vector Machine** | 0.872<br>[0.855, 0.890] | 0.945<br>[0.934, 0.956] | 0.853<br>[0.835, 0.872] | 0.887<br>[0.870, 0.904] | 0.870<br>[0.850, 0.888] |
| **Gradient Boost Machine** | 0.894<br>[0.878, 0.910] | 0.963<br>[0.955, 0.972] | 0.878<br>[0.861, 0.895] | 0.907<br>[0.892, 0.922] | 0.892<br>[0.875, 0.910] |
| **Multi-Layer Perceptron** | 0.860<br>[0.842, 0.878] | 0.932<br>[0.919, 0.945] | 0.864<br>[0.845, 0.882] | 0.857<br>[0.839, 0.876] | 0.860<br>[0.840, 0.880] |
| **Long Short-Term Memory Neural Network** | 0.874<br>[0.856, 0.891] | 0.927<br>[0.913, 0.941] | 0.875<br>[0.858, 0.893] | 0.873<br>[0.855, 0.89] | 0.874<br>[0.855, 0.892] |

**Figure 3** presents the AUC-ROC and Precision-Recall curves for each model using the top 30 input features. GBM achieved the highest AUROC (0.96), followed by LR and RF (0.95), and SVM (0.94). In terms of Precision-Recall curves, GBM also led with an area under the curve (AUC) of 0.97, while LR, RF, and SVM followed closely at 0.95, and MLP and LSTM at 0.93. These results reinforce GBM’s superiority in predicting hypertension-related mortality, even with a reduced feature set.

### Comparative analysis

We conducted a comparative analysis between models trained on all available features and those trained on the top 30 selected features. The results demonstrate that reducing the number of input features from over 400 to just 30 did not significantly degrade performance for most models. This finding underscores the effectiveness of our feature selection process. Across all evaluation metrics—accuracy, sensitivity, and F1 score—every model outperformed the logistic regression (LR) baseline. However, regarding AUROC, the two deep learning models showed a slight decline in performance relative to the others. Notably, performance trends emerged when grouped by model type. Tree-based models such as Random Forest (RF) and Gradient Boosting Machine (GBM) exhibited a stronger ability to capture complex patterns associated with hypertension-related mortality while simultaneously mitigating overfitting. This advantage is likely attributable to their hierarchical structure and ensemble learning strategy. These trends were consistent across both five-fold cross-validation and test set evaluations. Among all models, GBM achieved the highest performance across most metrics, surpassing even its tree-based counterpart, RF. This highlights GBM’s robust capability to distinguish between patients who survived and those who did not. RF also demonstrated substantial improvements over the baseline, ranking second among the six models evaluated.

### Model Explainability

To interpret model predictions, we calculated the mean absolute SHAP values across all six trained models, as shown in **Figure 4**. This analysis identified the top 30 features most predictive of hypertension-related mortality. Among these, 21 were laboratory variables (e.g., GCS - Eye Opening, Braden Mobility, Urea Nitrogen), 4 were related to admission type (e.g., Emergency, Urgent), 3 were demographic (e.g., anchor age, ICU stay, hospital stay length), 1 was medication-related (5% Dextrose), and 1 was a complication feature (Pneumonia).

**Figure 4.**
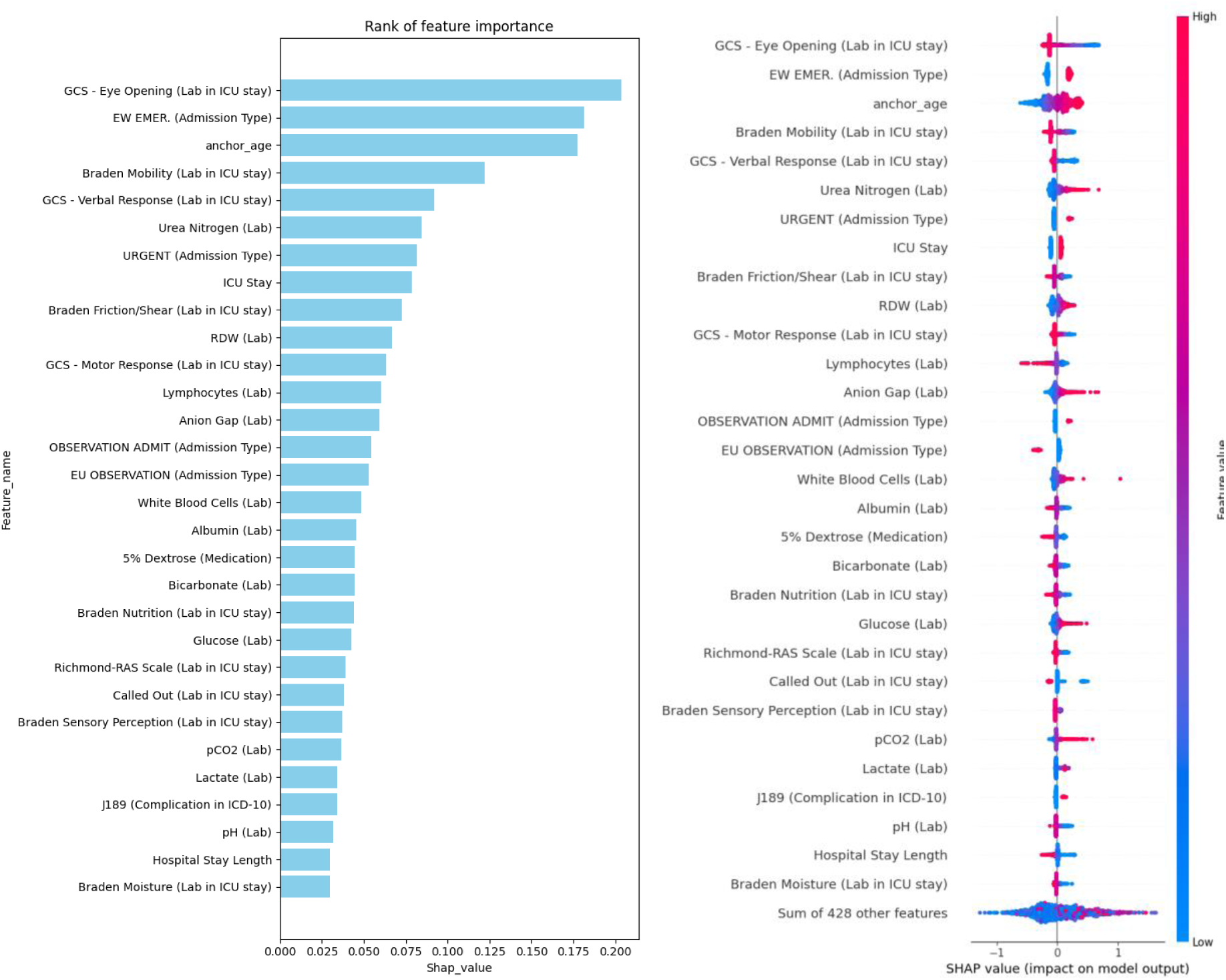
Ranking of the top 30 features based on average SHAP values, with information sources specified in the feature names.

Interestingly, several top-ranked features belong to specific clinical assessment tools. For example, the Glasgow Coma Scale (GCS), a standard measure of consciousness, was represented by all three components—eye opening, motor response, and verbal response— within the top 30 features, emphasizing its prognostic relevance. Another key group of features were derived from the Braden Scale (BS), used to assess pressure ulcer risk. Five distinct BS sub-scores appeared among the top features: Braden Mobility, Friction/Shear, Nutrition, Sensory Perception, and Moisture. Additional important laboratory markers included RDW, lymphocyte count, anion gap, white blood cell count, and albumin. For local interpretability, we selected two representative cases from the hypertension cohort and applied SHAP to explain individual predictions using the GBM model—the best-performing model. One case resulted in a positive prediction (mortality), and the other negative (non-mortality). Only features with SHAP contribution scores above the default 0.05 threshold are visualized. In the positive case (**Supplementary Figure 1**), key contributors to the high predicted mortality risk included two GCS components, three Braden scores, and bicarbonate level. In the negative case (**Supplementary Figure 2**), despite some mortality-risk indicators (e.g., lymphocyte count and urea nitrogen), protective features—such as non-urgent admission type, Braden Mobility score, and RDW—contributed more strongly to a negative prediction. These case studies reinforce the overall significance of our top 30 features in predicting hypertension-related mortality.

### Fairness evaluation

We further assessed model fairness using several debiasing techniques and compared results across models trained on all features versus the top 30. Table 4 presents the Disparate Parity Difference (DPD) and Equal Opportunity Difference (EOD) metrics with 95% confidence intervals for each model under different conditions. The Long Short-Term Memory (LSTM) model consistently demonstrated the best fairness across all scenarios. Without any debiasing, the LSTM achieved a DPD of 0.015 (95% CI: 0.001, 0.068) using all features, and 0.004 (95% CI: 0.001, 0.061) with the top 30 features. Its EOD values were similarly low: 0.008 (95% CI: 0.005, 0.066) for all features and 0.004 (95% CI: 0.005, 0.065) for the reduced feature set.

**Table 4.** Fairness evaluation of models based on demographic parity difference (DPD) and equalized odds difference (EOD).

| Metric | All features |  |  |  |  |  |  |  | Top 30 features |  |  |  |  |  |  |  |
| --- | --- | --- | --- | --- | --- | --- | --- | --- | --- | --- | --- | --- | --- | --- | --- | --- |
|  | DPD |  |  |  | EOD |  |  |  | DPD |  |  |  | EOD |  |  |  |
| Models \ Methods | No debiasing, 95%CI | Correlation Removal, 95%CI | Reduction, 95%CI | Threshold Optimization, 95%CI | No debiasing, 95%CI | Correlation Removal, 95%CI | Reduction, 95%CI | Threshold Optimization, 95%CI | No debiasing, 95%CI | Correlation Removal, 95%CI | Reduction, 95%CI | Threshold Optimization, 95%CI | No debiasing, 95%CI | Correlation Removal, 95%CI | Reduction, 95%CI | Threshold Optimization, 95%CI |
| **Logistic Regression** | 0.032<br>[0.002, 0.084] | 0.028<br>[0.002, 0.081] | 0.032<br>[0.002, 0.083] | 0.021<br>[0.001, 0.073] | 0.044<br>[0.011, 0.096] | 0.038<br>[0.010, 0.086] | 0.044<br>[0.011, 0.097] | 0.028<br>[0.007, 0.080] | 0.018<br>[0.001, 0.071] | 0.018<br>[0.001, 0.073] | 0.018<br>[0.001, 0.071] | 0.008<br>[0.001, 0.062] | 0.028<br>[0.007, 0.079] | 0.028<br>[0.007, 0.082] | 0.028<br>[0.007, 0.084] | 0.006<br>[0.005, 0.068] |
| **Random Forest** | 0.022<br>[0.001, 0.076] | 0.022<br>[0.001, 0.071] | 0.022<br>[0.001, 0.072] | 0.066<br>[0.015, 0.119] | 0.016<br>[0.006, 0.067] | 0.029<br>[0.007, 0.082] | 0.016<br>[0.005, 0.069] | 0.114<br>[0.055, 0.174] | 0.005<br>[0.001, 0.065] | 0.016<br>[0.001, 0.068] | 0.005<br>[0.001, 0.061] | 0.067<br>[0.013, 0.117] | 0.014<br>[0.005, 0.067] | 0.020<br>[0.006, 0.067] | 0.014<br>[0.006, 0.069] | 0.077<br>[0.048, 0.131] |
| **Support Vector Machine** | 0.032<br>[0.002, 0.080] | 0.026<br>[0.002, 0.081] | 0.002<br>[0.001, 0.060] | 0.014<br>[0.001, 0.066] | 0.055<br>[0.012, 0.104] | 0.049<br>[0.013, 0.100] | 0.036<br>[0.010, 0.086] | 0.032<br>[0.008, 0.080] | 0.012<br>[0.001, 0.066] | 0.012<br>[0.001, 0.07] | 0.015<br>[0.001, 0.069] | 0.006<br>[0.001, 0.063] | 0.019<br>[0.006, 0.077] | 0.019<br>[0.006, 0.074] | 0.025<br>[0.007, 0.078] | 0.022<br>[0.008, 0.079] |
| **Gradient Boost Machine** | 0.037<br>[0.002, 0.091] | 0.037<br>[0.002, 0.093] | 0.029<br>[0.002, 0.081] | 0.031<br>[0.002, 0.083] | 0.058<br>[0.019, 0.105] | 0.058<br>[0.018, 0.108] | 0.034<br>[0.008, 0.080] | 0.048<br>[0.012, 0.097] | 0.029<br>[0.001, 0.082] | 0.029<br>[0.002, 0.084] | 0.024<br>[0.001, 0.077] | 0.025<br>[0.001, 0.077] | 0.036<br>[0.008, 0.085] | 0.036<br>[0.008, 0.091] | 0.033<br>[0.008, 0.086] | 0.034<br>[0.006, 0.085] |
| **Multi-Layer Perceptron** | 0.024<br>[0.002, 0.078] | 0.021<br>[0.001, 0.073] | 0.024<br>[0.002, 0.077] | 0.024<br>[0.002, 0.077] | 0.026<br>[0.007, 0.072] | 0.026<br>[0.008, 0.075] | 0.026<br>[0.007, 0.075] | 0.026<br>[0.007, 0.075] | 0.020<br>[0.001, 0.070] | 0.017<br>[0.001, 0.068] | 0.020<br>[0.001, 0.075] | 0.020<br>[0.001, 0.071] | 0.044<br>[0.013, 0.097] | 0.031<br>[0.008, 0.083] | 0.044<br>[0.013, 0.096] | 0.027<br>[0.007, 0.076] |
| **Long Short-Term Memory Neural Network** | 0.015<br>[0.001, 0.068] | 0.014<br>[0.001, 0.067] | 0.015<br>[0.001, 0.068] | 0.012<br>[0.001, 0.062] | 0.008<br>[0.005, 0.066] | 0.016<br>[0.006, 0.069] | 0.008<br>[0.005, 0.063] | 0.006<br>[0.006, 0.064] | 0.004<br>[0.001, 0.061] | 0.006<br>[0.001, 0.061] | 0.004<br>[0.001, 0.059] | 0.013<br>[0.001, 0.065] | 0.004<br>[0.005, 0.065] | 0.017<br>[0.006, 0.075] | 0.004<br>[0.005, 0.063] | 0.006<br>[0.006, 0.068] |

In contrast, the GBM model exhibited the poorest fairness performance, with DPD values of 0.037 (all features) and 0.029 (top 30), and EOD values of 0.058 (all features) and 0.036 (top 30). Debiasing strategies yielded measurable improvements. For example, correlation removal reduced LR’s DPD from 0.032 to 0.028 and SVM’s from 0.032 to 0.026. The reweighting method improved both DPD and EOD for SVM and GBM, while threshold optimization proved effective for most models, except RF. These findings highlight the potential of targeted mitigation techniques to enhance algorithmic fairness without sacrificing predictive performance.

## DISCUSSION

### Interpretation of Findings

In this study, we developed and evaluated four machine learning models (Logistic Regression, Support Vector Machine, Random Forest, and Gradient Boosting Machine) and two deep learning models (Multilayer Perceptron and Long Short-Term Memory) to predict mortality among patients with hypertension using the MIMIC-IV dataset. By extracting a comprehensive set of features—including clinical, demographic, laboratory, medication, and complication-related variables—we aimed to enhance model performance and support clinical decision-making. Our findings demonstrate that the Gradient Boosting Machine (GBM) outperformed other models in terms of accuracy, sensitivity, F1 score, and AUROC. This suggests GBM’s superior capability to model the complex, non-linear relationships inherent in clinical data. Notably, tree-based models like GBM and Random Forest also showed resilience against overfitting and were more effective in capturing patterns related to hypertension mortality. These models’ ensemble learning strategies and hierarchical structure may explain their stronger performance relative to traditional methods and even deep learning models in this context.[18]

Importantly, we found that reducing the feature set from over 400 variables to the top 30 features, based on SHAP importance scores, did not significantly degrade model performance. This reinforces the value of our feature selection process and highlights the potential for streamlined, interpretable, and computationally efficient models that could be implemented in real-world clinical settings using EHR data similar to MIMIC-IV. The most predictive features identified, such as Glasgow Coma Scale (GCS) scores, Braden Scale scores, blood urea nitrogen, age, and complications like pneumonia, align well with clinical intuition and prior studies.[19, 20] For instance, elevated blood urea nitrogen has been shown to predict adverse cardiovascular outcomes, and age is a well-established risk factor for hypertension-related mortality.[21] Red Cell Distribution Width (RDW) has emerged in recent literature as a marker for mortality in cardiovascular diseases, while variables like lymphocyte count, bicarbonate, and lactate have also been linked to in-hospital mortality in hypertensive or critically ill populations.[22, 23] Our findings further emphasize the prognostic importance of neurological and functional assessments such as the GCS and Braden Scale. Low GCS scores—particularly in eye, motor, and verbal responses—were strongly associated with higher mortality, consistent with literature on traumatic brain injury and critical illness.[24] Similarly, five Braden sub-scores were negatively associated with mortality risk, echoing findings from COVID-19 mortality studies that associated lower Braden scores with worse outcomes.[25]

### Fairness Evaluation

We also examined model fairness through the lens of two standard metrics: Demographic Parity Difference (DPD) and Equalized Odds Difference (EOD). To mitigate potential biases, we applied three debiasing strategies—correlation removal, reduction, and threshold optimization. Overall, the Long Short-Term Memory (LSTM) model demonstrated the best fairness across scenarios, achieving the lowest DPD and EOD scores, especially when trained on the top 30 features. Interestingly, models trained on the reduced feature set generally exhibited better fairness metrics compared to those using all features, suggesting that the feature selection process may inherently reduce biases. While the debiasing methods improved fairness for most models trained on the full feature set, they were less impactful when applied to models using the top 30 features. This pattern suggests that reducing the feature space to the most important variables may inherently support more equitable predictions—potentially by minimizing noise or spurious correlations linked to sensitive attributes.

### Comparison with Existing Models

Compared to prior studies in hypertensive patient risk stratification, our work offers several advancements. First, we leveraged a broader and more comprehensive set of features extracted from the MIMIC-IV dataset, including novel variables such as ICU length of stay and specific complications. Second, we systematically compared multiple machine learning and deep learning models, rather than focusing on a single approach. Third, we incorporated a robust model interpretability framework using SHAP values and conducted an in-depth fairness evaluation—an area often neglected in earlier studies. Moreover, our comparison of models trained on all features versus only the top 30 features revealed minimal performance trade-offs. This not only validates the importance of the identified top predictors but also offers a path toward more efficient and interpretable clinical models. Reducing the feature set can ease the burden of data collection and processing in clinical environments, making deployment more feasible across various EHR systems.

### Limitations

Despite the promising results, this study has several limitations that should be acknowledged. First, our analysis was based solely on data from the MIMIC-IV database, which, while comprehensive, represents a single healthcare system and may not generalize to other populations or clinical settings. External validation using data from different institutions, regions, or healthcare systems is necessary to confirm the robustness and generalizability of our models. Additionally, the exclusion of patients with incomplete records could introduce bias, as these patients may have distinct characteristics compared to those included in the study. Second, the definition of hypertension-related mortality was inferred from the available data and may not fully capture the complexity of clinical decision-making or cause of death in critically ill patients.[26] Further refinement in outcome definitions, including cause-specific mortality, could enhance model accuracy and relevance. Third, while we assessed and mitigated bias using fairness metrics and debiasing strategies, our analysis was limited to demographic parity and equalized odds. Other aspects of algorithmic fairness, such as calibration across subgroups or individual-level fairness, were not explored and warrant future investigation.[27] Finally, while we identified top predictive features and demonstrated their utility in building more efficient models, clinical validation of these features—and how they might be used in practice to guide treatment decisions—remains an area for future research.[28, 29] Integration into clinical workflows will also require collaboration with clinicians, careful usability testing, and assessments of real-world impact.[30, 31]

Our findings suggest that machine learning models, particularly GBM, can play a valuable role in predicting hypertension-related mortality and potentially guiding clinical decision-making in intensive care settings. The identification of interpretable and clinically relevant predictors, combined with fairness-aware modeling strategies, strengthens the potential for safe and equitable integration into clinical workflows.[32] Future work should explore the external validation of our models in different hospital systems and populations, as well as real-time deployment feasibility.[33] Further studies may also investigate causal relationships between identified features and outcomes, and assess how these models could support or augment clinician judgment at the bedside.

## CONCLUSION

This study developed and evaluated multiple machine learning and deep learning models to predict in-hospital mortality among patients with hypertension using the MIMIC-IV dataset. Our findings highlight the strong performance of the gradient boosting machine (GBM) model, which outperformed other approaches in capturing the complex, non-linear relationships inherent in clinical data. By incorporating a broad set of features, including novel variables related to patient complications and ICU stays, and by identifying a reduced set of the most important predictors, we demonstrated that accurate and efficient mortality prediction is achievable using routinely collected EHRs data. Additionally, we evaluated the fairness of our models across demographic subgroups and applied debiasing strategies to mitigate potential disparities in performance. Our results indicate that feature selection may play a role in enhancing model fairness, offering a promising direction for building more equitable clinical prediction tools. These findings support the potential of interpretable, fair, and efficient machine learning models to assist in clinical decision-making for critically ill patients with hypertension. Future work should focus on external validation, clinical integration, and real-world impact evaluation to ensure these models can meaningfully improve patient outcomes in diverse care settings.

## CONTRIBUTION STATEMENT

SZ, SD, and ZX contributed to data analyses and the writing of the manuscript. JY designed the study, contributed to the data analyses and the writing of the manuscript. All authors read and approved the final version of the manuscript.

## DATA AVAILABILITY STATEMENT

All data are available at Medical Information Mart for Intensive Care: https://mimic.physionet.org/. The relevant code and analyses are available at: https://github.com/ShenghanZhang1123/hypertension_mortality_pred.

## SUPPLEMENTARY MATERIAL

**Supplementary Table 1.**
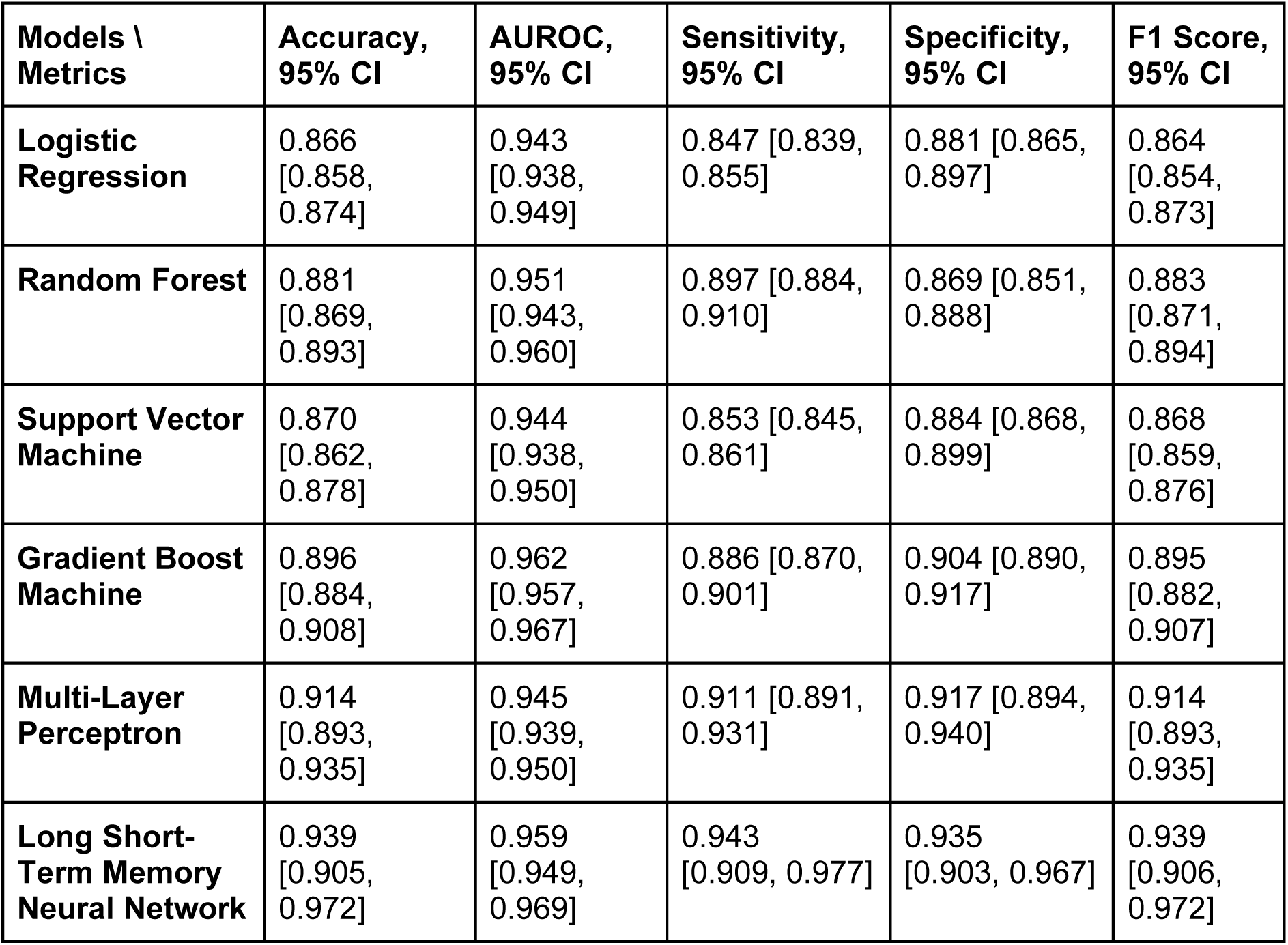
Summary of cross-validation results for model performance on the validation set in hypertension mortality prediction using all features.

**Supplementary Table 2.** Summary of cross-validation results for model performance on the validation set in hypertension mortality prediction using top 30 features.

| **Models \ Metrics** | **Accuracy** | **AUROC** | **Sensitivity** | **Specificity** | **F1 Score** |
| --- | --- | --- | --- | --- | --- |
| **Logistic Regression** | 0.861<br>[0.849, 0.873] | 0.938<br>[0.932, 0.945] | 0.840<br>[0.828, 0.852] | 0.876<br>[0.859, 0.894] | 0.858<br>[0.843, 0.872] |
| **Random Forest** | 0.875<br>[0.863, 0.887] | 0.946<br>[0.940, 0.952] | 0.882<br>[0.867, 0.897] | 0.869<br>[0.854, 0.884] | 0.876<br>[0.863, 0.889] |
| **Support Vector Machine** | 0.866<br>[0.851, 0.881] | 0.940<br>[0.934, 0.947] | 0.849<br>[0.838, 0.861] | 0.879<br>[0.857, 0.901] | 0.864<br>[0.847, 0.880] |
| **Gradient Boost Machine** | 0.892<br>[0.884, 0.899] | 0.960<br>[0.956, 0.964] | 0.886<br>[0.873, 0.898] | 0.897<br>[0.886, 0.907] | 0.891<br>[0.882, 0.900] |
| **Multi-Layer Perceptron** | 0.871<br>[0.863, 0.880] | 0.932<br>[0.927, 0.938] | 0.868<br>[0.864, 0.872] | 0.873<br>[0.858, 0.889] | 0.871<br>[0.862, 0.879] |
| **Long Short-Term Memory Neural Network** | 0.881<br>[0.862, 0.899] | 0.935<br>[0.926, 0.944] | 0.889<br>[0.875, 0.903] | 0.875<br>[0.851, 0.898] | 0.882<br>[0.863, 0.900] |

**Supplementary Figure 1.**
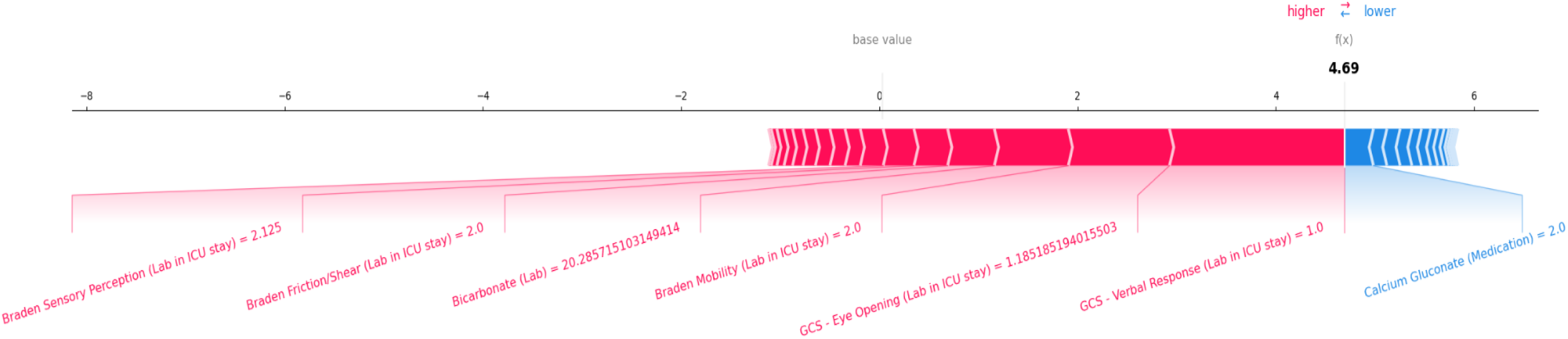
Local explanation example for the positive (mortality) case.

**Supplementary Figure 2.**
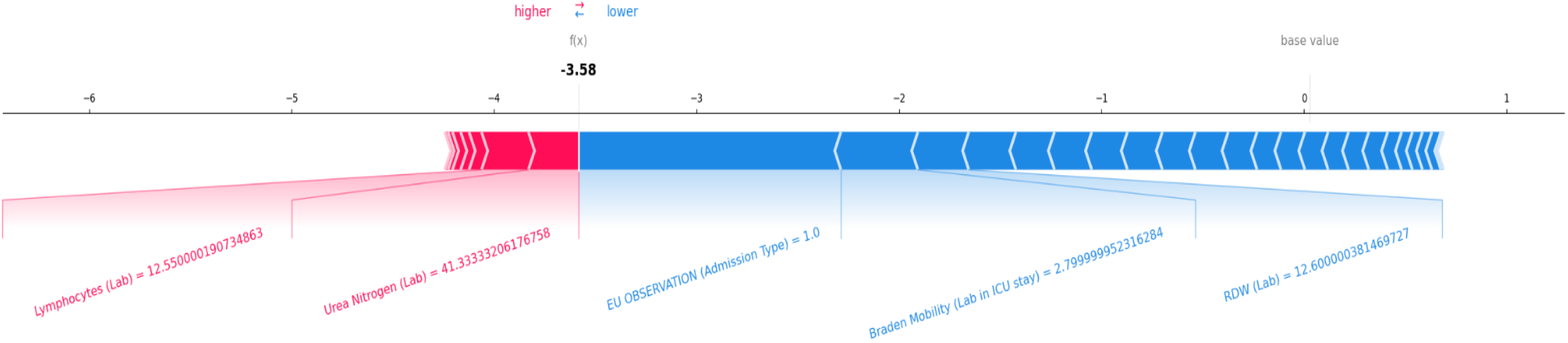
Local explanation example for the negative (non-mortality) case.

